# Optical genome mapping as a next-generation cytogenomic tool for detection of structural and copy number variations for prenatal genomic analyses

**DOI:** 10.1101/2021.02.19.21251714

**Authors:** Nikhil Shri Sahajpal, Hayk Barseghyan, Ravindra Kolhe, Alex Hastie, Alka Chaubey

**Author notes:** Corresponding author: Alka Chaubey, PhD, FACMG, Chief Medical Officer, Bionano Genomics, Inc., 9540 Towne Centre Drive, Suite 100, San Diego, CA 92121, E, C. Equal contribution authors (Nikhil Shri Sahajpal, Hayk Barseghyan).

## Abstract

Global medical associations (ACOG, ISUOG, ACMG) recommend diagnostic prenatal testing for the detection and prevention of genetic disorders. Historically, cytogenetic methods such as karyotype analysis, fluorescent in situ hybridization (FISH), and chromosomal microarray (CMA) are utilized worldwide to diagnose common syndromes. However, the limitations of each of these methods, either performed in tandem or simultaneously, demonstrates the need of a revolutionary technology that can alleviate the need of multiple technologies. Optical genome mapping (OGM) is a novel technology that fills this void by being able to detect all classes of structural variations (SVs), including copy number variations (CNVs). OGM is being adopted by laboratories as a next-generation cytogenomic tool for both postnatal constitutional genetic disorders and hematological malignancies. This commentary highlights the potential of OGM to become a standard of care in prenatal genetic testing by its ability to identify large balanced and unbalanced SVs (currently the strength of karyotyping and metaphase FISH), CNVs (by CMA), repeat contraction disorders (by Southern blotting) and multiple repeat expansion disorders (by PCR based methods or Southern blotting). Also, next-generation sequencing (NGS) methods are excellent at detecting sequence variants but are unable to accurately detect the repeat regions of the genome which limits the ability to detect all classes of SVs. Notably, multiple molecular methods are used to identify repeat expansion and contraction disorders in routine clinical laboratories around the world. With non-invasive prenatal screening test (NIPT) as the standard of care screening assay for all global pregnancies, we anticipate OGM as a high-resolution cytogenomic diagnostic tool employed following a positive NIPT screen or for high-risk pregnancies with an abnormal ultrasound. Accurate detection of all types of genetic disorders by OGM, such as liveborn aneuploidies, sex chromosome anomalies, microdeletion/microduplication syndromes, repeat expansion/contraction disorders is key to reducing the global burden of genetic disorders.

## Introduction

Global medical associations, such as American College of Obstetrics and Gynecology (ACOG), International Society of Ultrasound and Obstetrics and Gynecology (ISUOG), and American College of Medical Genetics and Genomics (ACMG), recommend prenatal genetic testing including non-invasive prenatal screening test (NIPT) and invasive diagnostic testing be offered to all pregnant women, irrespective of the gestation age and maternal age [1-3]. Typically, NIPT can be offered to pregnant women as early as ≤ 10 weeks of gestation [4]. Following a positive screen or a “no-call” result that may be due to technical limitations of each NIPT platform, invasive diagnostic testing is recommended to confirm the findings of the screening test. Also, pregnancies that have an abnormal ultrasound showing fetal defects are recommended to be followed up with diagnostic invasive testing, which includes chorionic villus sampling (CVS), amniocentesis or periumbilical blood sampling (PUBS) performed at either 10-14 weeks or >15 weeks of pregnancy [5].

Since the early 1980’s confirmatory diagnostic testing included conventional cytogenetic methods such as karyotyping and FISH [6,7]. In the 2000’s, chromosomal microarray analysis (CMA) using amniocytes or CVS to provide a comprehensive genetic profile in pregnancies with suspected fetal genetic disease was recommended by ACOG and ISUOG as the first line test in high-risk pregnancies with an abnormal ultrasound (8,9). More recently, fetal exome sequencing has been considered as a diagnostic test to identify the genetic cause of fetal genetic disorders [8]. However, the complete profiling of amniocytes or CVS using these multiple technologies, in a tiered fashion, or simultaneously, is time consuming and cost prohibitive. The sensitivity and specificity of karyotyping can affect reporting, as structural variant (SV) locations and sizes remain inaccurate and some translocations remain cryptic. FISH panels require *a priori* knowledge and it is difficult to detect all common microdeletions and microduplications. CMA, despite having higher resolution for the detection of copy number variants (CNVs) is limited in its ability to detect balanced translocations, inversions and low-level mosaicism [10]. Fetal exome sequencing can identify sequence variants and some copy number aberrations, but the sequencing technology bias does not allow for the detection of several classes of large SVs [8].

For the past several decades, cytogenomics has been at a standstill in prenatal diagnostics and there exists a need for a disruptive, novel, and high-resolution technology that can detect clinically significant SVs in a single assay. Optical genome mapping (OGM) has been recognized as a key genomic technology for the detection of all classes of SVs in many disorders [11-13]. OGM has been extensively utilized to characterize SVs in postnatal and hematologic diagnostics, demonstrating a 100% clinical concordance with traditional cytogenetic analysis, and identifying additional clinically relevant abnormalities that remained beyond the purview of current technologies [11,12,14,15]. Recently, OGM using Saphyr® demonstrated a 100% clinical concordance compared to combined cytogenetic analysis for the detection of 100 abnormalities in a cohort of 85 patients with constitutional disorders that included several sample types such as amniocytes, CVS and lymphoblastoid cells [11]. The study included 34 microdeletions or duplications, 7 aneuploidies, 28 balanced translocations as well as ring chromosomes, inversions, and complex rearrangements. This diverse set of abnormalities represents a good selection of SV classes and serves as a good foundation for exploring more difficult variations and mixtures. Another study has done just that, Shieh et al [12], have attempted to find genetic diagnoses for 50 cases of individuals with developmental delay or intellectual disability where the standard of care had failed to provide a definitive diagnosis. Within this cohort, they were able to find pathogenic or likely pathogenic variations in 12% of cases using OGM.

Prenatal Fragile X testing may be recommended if there is a history of fragile X in a family or if the mother is a carrier. In such cases, a specialized test is performed, sometimes in conjunction with other diagnostic tests. Supporting the ability to effectively assay for repeat expansions, Otero et al. have measured repeat expansion in DM1 and DMPK from patients with myotonic dystrophy using OGM and were able to show that the repeat length correlated with the degree of mRNA splicing and severity of symptoms, these repeats ranged up to 15kbp [16].

Three studies on OGM technology have recently demonstrated its ability to identify multiple genomic aberrations associated with hematologic malignancies. The first study aimed to detect all clinically relevant SVs reported by multiple methods including karyotyping, CMA, and FISH, in leukemias from bone marrow or peripheral blood from 48 cases. The authors were able to detect all SVs and CNVs that were above 10% allele fraction in all 48 cases including several different IGH fusions, *BCR-ABL1* translocations, etc [17]. The second study focused on acute myelogenous leukemia (AML) in 100 cases, and the authors were also able to reproduce all clinically reported genomic abnormalities [14]. This study also demonstrated the identification of SVs deemed clinically relevant according to European Leukemia-Net (ELN) guidelines [18] in an additional 11% of cases. The third study aimed to comprehensively define SVs in myelodysplastic syndromes (MDS) [15]. In this study, the authors reported detection of all clinically actionable genomic aberrations that were above the limit of detection and found additional SVs in 33% of patients, which were later confirmed by orthogonal methods.

Clearly, multiple studies have demonstrated the performance of OGM and its ability to stand out as a unique technology for the detection of all classes of clinically significant genome-wide SVs. Here, we present the methodology and the application of OGM in the prenatal setting, with classical examples of several syndromic SVs that are important for the diagnosis and detection of prenatal genetic disorders.

## Material and Methods

### Prenatal workflow

Optical genome mapping using Bionano Genomics Saphyr prenatal workflow has been optimized and tailored to utilize either direct or cultured amniotic fluid cells or chorionic villus samples (CVS) as in clinical practice for current standard of care methods such as karyotyping, FISH or CMA. If the amniotic fluid does not contain 1.5 million cells for proceeding with DNA extractions, it is recommended to perform cell culturing. Similarly, if the microscopic dissections of the CVS do not yield enough starting material, cell culturing is recommended. In this study, amniocyte cultures or lymphoblastoid cell lines were evaluated. First, cells were dislodged by gentle pipetting when they reached approximately 80-90% confluency. Cells were pelleted by centrifugation and resuspended in a cell buffer (Bionano Genomics, Inc., San Diego, CA, USA) for ultra-high molecular weight (UHMW) DNA extraction via the Bionano Prep SP DNA Isolation Kit. Subsequently, the Bionano Prep DLS Labeling Kit was used to fluorescently label long molecules at specific sequence motifs throughout the genome. The labeled DNA was loaded onto Saphyr chips for linearization and imaging in massively parallel nanochannel arrays. The observed unique patterns on single long DNA molecules were used for *de novo* genome assembly and structural variant calling via the Bionano Solve pipeline (version 3.6) (Figure 1). The study was approved by the IRB A-BIOMEDICAL I (IRB REGISTRATION #00000150), Augusta University. HAC IRB # 611298. Based on the IRB approval, all PHI was removed and all data was anonymized before accessing for the study.

**Figure 1.**
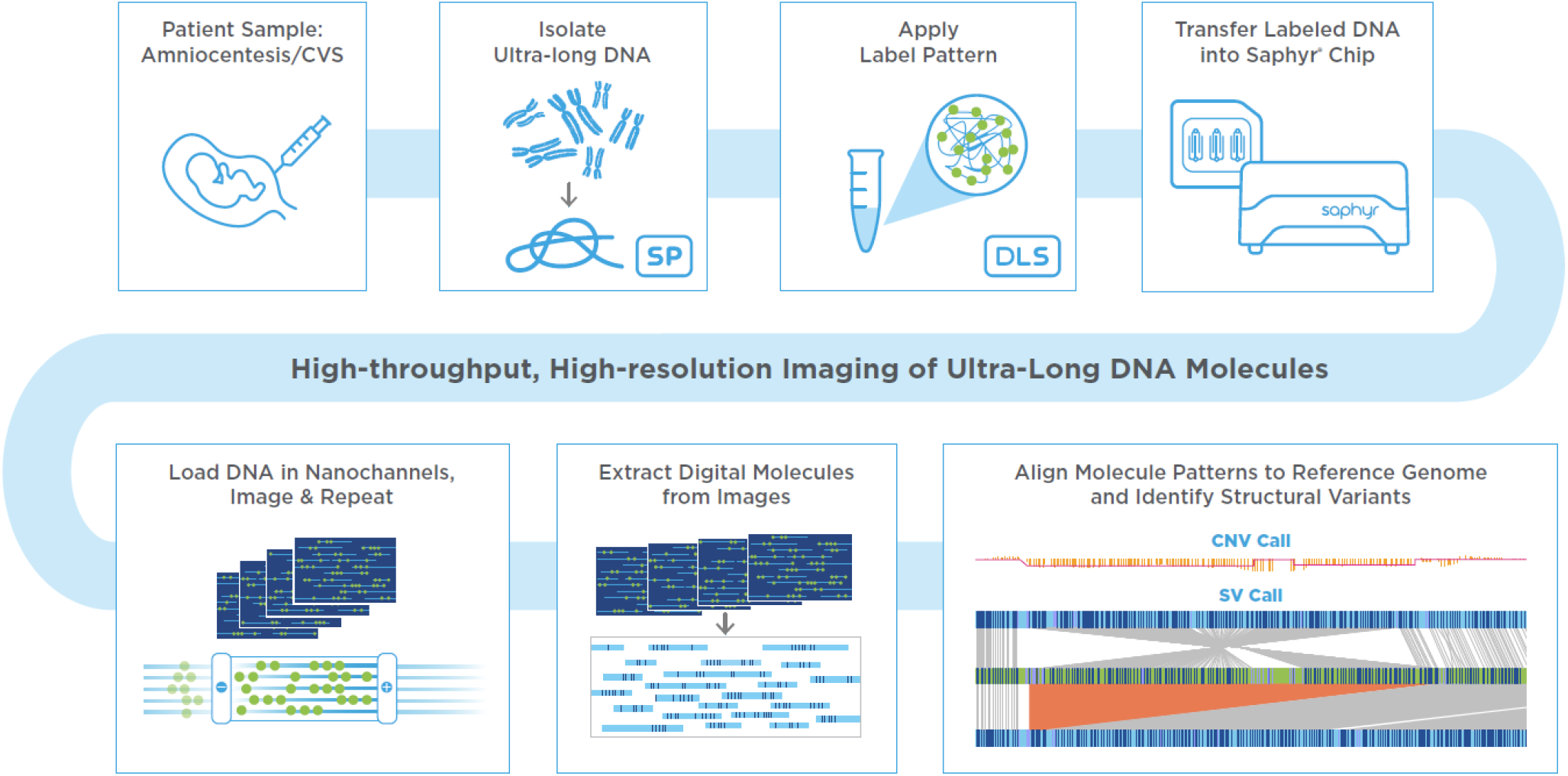
Prenatal workflow for optical genome mapping From top left to bottom right: Sample format can be from 1 million cultured cells, directly from cells contained in amniotic fluid or CVS sample and can be fresh or frozen. DNA is subsequently labelled at a 6 bp motif by the DLS labeling technology creating a label pattern which is unique throughout th genome. Labelled DNA is then loaded on a Saphyr Chip where DNA molecules are linearized and imaged in repeated cycles. Images are processed to extract molecules that contain the linear positions of sequence motif labels. Multiple molecules are used to create consensus genome maps representing different alleles from the sample. The sample’s optical genome map is aligned to the reference genome and differences are automatically called allowing for detection of structural variations in a genome wid fashion.

### Assay Quality Control

The OGM protocol has several quality control (QC) metrics, at both pre-analytical and analytical stages. The pre-analytical QC metrics include the presence of UHMW DNA – observable viscosity/clarity of DNA during pipetting and a minimum DNA concentration of >35 ng/µl needed for subsequent labeling. The analytical QC metrics include label density of ∼15/100 kbp, average filtered N50 > 230 kbp, map rate > 70%, and effective coverage of >80x for the generation of a *de-novo* assembly. All cultured amniocytes met these QC metrics, indicating that the Bionano Genomics prenatal workflow can generate high quality data similar to other already established workflows.

### Data Analysis

Bionano Access (version 1.6), an OGM specific structural variant analysis software available with a standard web browser application, links to bioinformatic servers running Bionano Solve (version 3.6), an automated analytical pipeline for detection of genomic abnormalities, used for data processing. Briefly, single molecules were used to generate *de novo* assembly of the human genome. Direct alignment of *de novo* maps as well as single molecules to the reference genome allowed identification of SVs, CNVs as well as aneuploidies. Table 1 shows a full list of detectable variant classes via Bionano Solve.

**Table 1.** Specification of optical genome mapping for the detection of different variant classes.

| Variant | Variant types |  | Variant Description | Bionano OGM |
| --- | --- | --- | --- | --- |
| Aneuploidy | Monosomy |  | Chromosome loss | ✓ |
|  | Trisomy |  | Chromosome gain | ✓ |
|  | Triploidy |  | Whole genome Triploidy | Not currently |
|  | Tetraploidy |  | Whole genome Tetraploidy | Not currently |
|  | Ring chromosome |  | CNV and fusion | ≥ 500 kbp + fusion break |
| Structural Variants | Copy Number Variants | Deletions/<br>Duplications | Interstitial | ≥ 500 bp |
|  |  |  | Terminal | ≥ 500 kbp |
|  |  | Insertions | Interstitial (unknown sequence) | ≥ 500 bp |
|  | Translocations |  | Balanced | ✓ |
|  |  |  | Unbalanced | ✓ |
|  | Inversions |  | Pericentric | ✓ |
|  |  |  | Paracentric | ≥ 30 kbp |
| Regions of Homozygosity | ROH |  | ROH | In development |
| Macrosatellite/<br>microsatellite | Repeats |  | Contractions/expansions | ≥ 500 bp |
| Single Nucleotide Variants | Small variants |  | SNVs, INDELs | – |

Bionano Access has numerous, built in, variant filtration options geared towards expedited identification of pathogenic genetic aberrations. Specifically, one of the most useful features is the custom-built database of SV consisting of >300 healthy individuals that allows a fast and efficient way to filter common, likely-benign variants and generating a list of rare, potentially deleterious SVs. Additionally, users can investigate variants overlapping with specific disease-causing genes or genetic loci via a specified user generated or already available gene lists. Other useful filtration criteria include SV confidence, size and SV supporting molecule cutoffs (i.e., number of molecules confirming the identified SV). This process generally results in a small list of SVs requiring an additional review for pathogenicity classification.

### OGM Turnaround Time

Bionano Genomics OGM workflow is optimized to deliver fast turnaround times. UHMW DNA extraction, molecule labeling and instrument run can all be accomplished in 3 days. As in standard cytogenomic procedures in a prenatal workflow, extra time may be allocated for cell culture growth. Automated data analysis pipelines with scalable cloud computing support, web interface and variant filtrations settings allow for fast processing for raw data into an actionable report.

## Results and Discussion

Screening for open neural tube defects and chromosomal aberrations is an important part of prenatal care. NIPT as a global screening test has changed the prenatal testing landscape dramatically in the past decade. The American College of Obstetricians and Gynecologists (ACOG) recently recommended that prenatal aneuploidy screening should be offered to all pregnant women regardless of age or known risk factors, compared to the previous recommendation where the screening for chromosomal anomalies was offered only to women >35 years, or with previous history of miscarriages [1].

The global incidence of congenital disorders is ∼6% (7.9 million infants) of which 50% of birth defects remain of unknown genetic cause [19]. The chromosomal aberrations that include large duplications, deletions of chromosomal segments or entire chromosomes can be determined during prenatal care. The liveborn trisomic syndromes such as, trisomy13 (Patau syndrome), 18 (Edwards syndrome) and 21 (Down syndrome) account for some of the most common birth defects [20]. This also set the basis of NIPT being the 1st tier screening test for these autosomal liveborn trisomies (trisomy 13, 18 and 21) and other sex chromosomal aneuploidies (Turner syndrome, Klinefelter syndrome, etc.) [1-3]. Not only must these NIPT screens be confirmed with invasive diagnostic testing (on amnio and CVS samples) currently performed by karyotyping, FISH and CMA, but other genomic aberrations are detected by invasive testing (beyond the scope of NIPT assays as a genome-wide screen). Taken together, these technologies are time consuming, limited in resolution, and are cost-prohibitive. The advent of Bionano OGM technology in clinical diagnostics is revolutionizing the practice of cytogenetics, and is becoming a next-generation cytogenomic tool that has the potential to replace the traditional cytogenetic methods in the coming years. In this commentary, we present a few examples of unique SV classes that have been identified by OGM (Figure 2).

**Figure 2.**
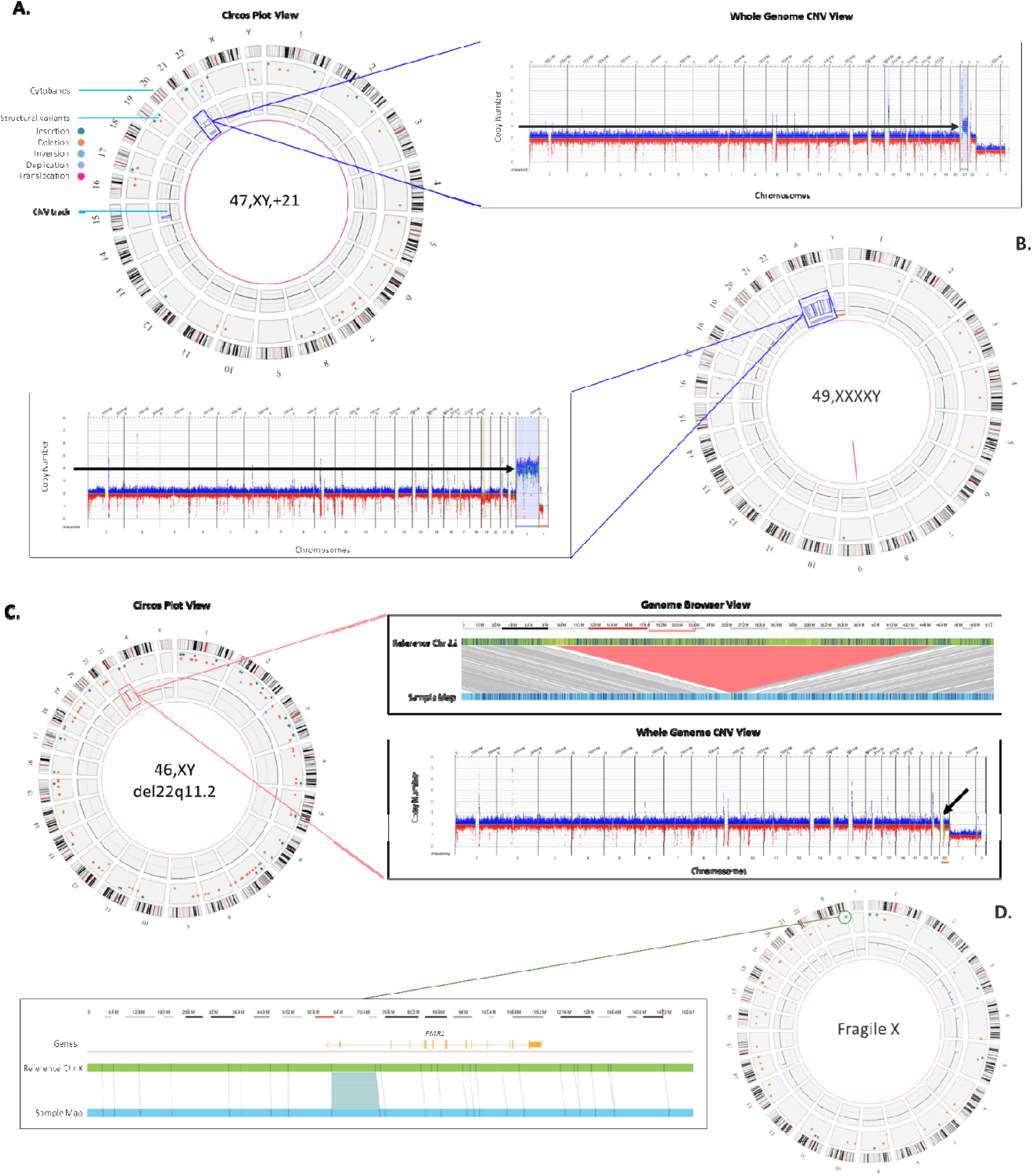
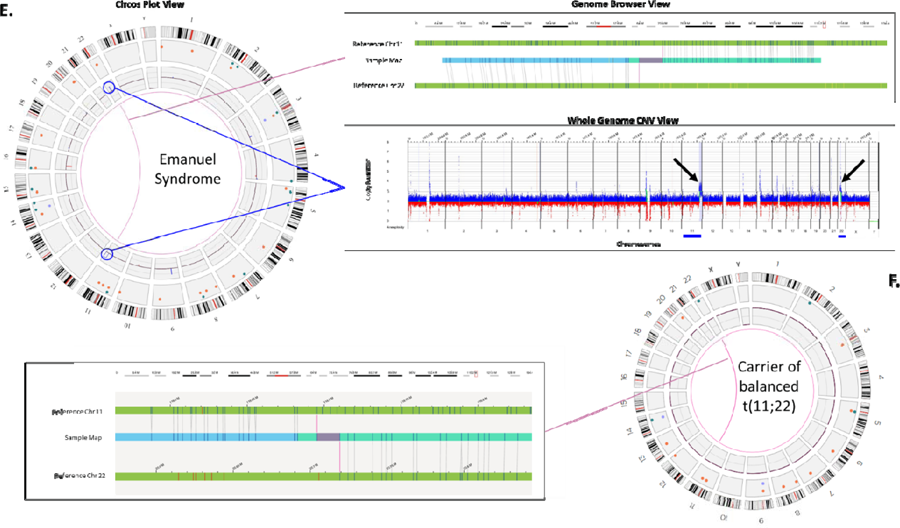
Examples of SVs identified by OGM **A:** Left panel: OGM circos plots are automatically generated and the default view is composed of the following layers: the outer circle displays cytoband locations, the middle circle displays color coded interstitial SVs that were identified in those particular locations, and the innermost circle displays observed copy number changes for each chromosome or region. Translocations are reported as lines in the center connecting the genomic loci involved (Figure 2E). **A:** a blue box on the circos plot around th inner circle of the CNV plot highlights the chromosome 21 gain, the right panel shows the whole genome CNV profile, a linear visualization of the CNV changes across the genome. The Y-axis represents the copy number change and X-axis lists the chromosome numbers. Gains are highlighted in blue while losses are highlighted in red. Here, the black arrow points to chromosome 21 which has 3 copies. **B:** Right panel: shows the circos plot summary displaying SVs and an aneuploidy in the sample. The blue box around the inner circle of the CNV plot points to chromosome X gain. Left panel: the CNV plot shows a gain of chromosome X. The black arrow points to chromosome X which is present in four copies. **C:** Left panel: Circos plot summary displaying SVs in the sample. The orange box around a region on chromosome 22 highlights a pathogenic deletion. Top right panel: The genome browser view details the alignment of the sample’s consensus map (light blue bar) with the reference consensus maps (light green bars) and provides the detail of the structural variation. Here the sample’s map alignment to the reference maps of chromosomes 22 illustrates a large ∼3Mbp deletion (light red). Bottom right panel: CNV plot showing loss on chromosome 22 (black arrow). **D:** Right panel: Circos plot summary displaying SVs in the sample. The green circle in the middle circle highlights an insertion identified on chromosome X. Left panel: The genome browser view details the alignment of the sample’s consensus map (light blue bar) with the reference chromosome X (light green bars) showing a highlighted region on the sample map that contains an insertion. The insertion is within the *FMR1* gene, inferred (and confirmed) to be a triplet repeat expansion. **E:** Left panel: Circos plot summary displaying SVs in the sample. Blue lines point to regions on chromosomes 11 and 22 with CNV gains. The purple line points to a translocation also observed between chromosomes 11 and 22. Top right panel: The genome browser view detailing the alignment of the sample’s consensus map (light blue bar) with the reference chromosome 11 and 22. Here the sample’s map aligns to two reference chromosomes indicates translocation. Bottom right panel: CNV plot showing CNV gains on chromosomes 11 and 22 (black arrows). **F:** The carrier mother of the case in Figure 2E showing a balanced translocation between chromosomes 11 and 22, but no CNV gains on either chromosome 11 or 22.

### Autosomal trisomy

Trisomy 21, or Down syndrome is the most common live-born trisomy syndrome [20]. Soft ultrasonography markers allude to the presence of trisomy 21 and liveborn babies have distinct facial and physical features, with congenital malformations including cardiovascular system, gastrointestinal tract and immune system [21]. The example in figure 2A shows the genome data in a circos plot as well as the copy number plot showing a gain of chromosome 21 identified with the OGM prenatal workflow.

### Sex Chromosome Aneuploidy

Disorders of sexual development (DSD), which include sex chromosome aneuploidies affect approximately 0.5% of the global population. The range of phenotypes for cases affected with DSD ranges from mild non-syndromic forms such as hypospadias to severe syndromic forms with complete sex reversal [22]. Figure 2B shows an example of a sex-chromosome aneuploidy case identified by OGM. Four copies of the X chromosome are present in the patient as seen by the circos plot on the left and linear representation of chromosomal copy number changes on the right.

### Microdeletion syndrome

DiGeorge syndrome is the most common microdeletion syndrome affecting approximately 1:4000 births, requiring preventive measures at birth. It occurs as a result of a ∼ 3Mb deletion at Chr 22q11.2 and the clinical symptoms include congenital heart defects, developmental delay, and hypocalcemia among other symptoms [23]. Figure 2C shows an example of the classical 2.87 Mb deletion of Chr 22q11.2. This deletion can be seen by the two complementary calling algorithms: the CNV profile provides dosage information and can be seen as a red dip in the circos plot (boxed) and also in the whole genome CNV display (arrow). The fusion for genomic DNA adjacent to the deletion provides further evidence of the deletion and gives high accuracy of the deletion breakpoints.

### Repeat expansion disorders

Certain triplet, tetra-, penta-, and hexa-nucleotide sequences can expand for thousands of base pairs resulting into pathogenic phenotypes such as fragile X, amyotrophic lateral sclerosis (ALS), and myotonic dystrophy [24]. Accurately detecting and measuring these repeat expansions is difficult since long tandem repeat measurement is refractory to molecular methods, yet too small to be detected by cytogenetic methods. Figure 2D shows the repeat expansion in the *FMR1* gene causing fragile X syndrome. The measurement of the size of the DNA fragment between two flanking labels is precise, within ∼ 60 base-pairs, or 20 repeat units. Therefore, an expansion above ∼>220 repeats could be utilized to “rule-in” as a pathogenic finding. In the example shown, the expansion is 867 bp or 289 triplet repeats. OGM can effectively measure repeat expansions that are at least 500 bp and up to 100 kbp in size.

### Unbalanced translocations (in fetus) secondary to a balanced translocation carrier (parent)

Emanuel Syndrome is caused by a chromosomal imbalance secondary to a parental balanced translocation involving chromosomes 11 and 22. The unbalanced +der (22) results due to the malsegregation of one derivative chromosome resulting in 47 chromosomes. The extra chromosome leaves the affected individual with three copies of portions of chromosomes 11 and 22 [25]. Figure 2E shows the OGM result for an individual with Emanuel syndrome, in this case the copy increases can be seen on chromosome 11 and 22 in the circos plot (circled) and in the whole genome CNV plot (arrows). Furthermore, the fusion of chromosome 11 and 22 can be seen in the circos plot by the pink line connecting the two chromosomes in the center and a zoom-in of the fusion in the genome browser where a genome map from the case can be seen connecting the two chromosomes. The most frequent balanced translocation in the human population, t(11;22) [25] in the carrier parent is highlighted in Figure 2F. OGM is also able to detect the carrier status through direct detection of the inter-chromosomal fusion in copy number neutral cells (Figure 2F).

To demonstrate the inter-site reproducibility of the OGM prenatal workflow, 5 samples were replicated on the same samples in two different sites, by different instruments, and different operators (unpublished data) indicating the robustness of the assay/method. In this reproducibility study, all assays passed the QC metrics at both sites and all clinical aberrations were detected at both sites. Going forward, larger reproducibility studies will be needed to further establish the robustness. Nevertheless, since OGM is much more automated with regard to data processing and SV calling compared to traditional cytogenetics, it is more objective and is expected to have fewer subjective operator determined differences (as observed with skilled cytogenetic personnel in any clinical laboratory). This also significantly reduces the burden of highly trained experts for data interpretation.

This short communication demonstrates the power and potential of OGM in detecting the classical genomic abnormalities which are commonly found in prenatal testing. Combining structural variations, copy number variations and repeat expansions into a single assay is unprecedented for any genomic technology.

## Conclusions

Optical genome mapping, with its power to detect all classes of SVs, including CNVs, at a higher resolution than traditional cytogenetic methods can play a significant role in prenatal care and management as a next-generation cytogenomic tool. OGM can be employed as an invasive prenatal testing tool after a positive NIPT screen and for the detection of microdeletion/duplication syndromes implicated in high-risk pregnancies. OGM has shown 100% concordance with the current combinatorial cytogenetic assays in several studies aimed at investigating complex genetic disorders, constitutional disorders and liquid tumors, while detecting all of the different types of chromosomal anomalies including aneuploidies, large deletions/duplications, CNVs, balanced chromosomal events and complex chromosomal rearrangements. The detection of the entire spectrum of cytogenetic aberration in a single assay, with the capability to identify unique genomic abnormalities and better characterize the SVs, demonstrates the potential of OGM technology to replace traditional cytogenetic assays. Additionally, as several publications have shown, OGM has the potential to identify novel clinically significant genetic abnormalities thereby increasing the diagnostic yield not only in the clinic, but also in the prenatal setting.

To conclude, OGM technology is cost-effective, has a fast turn-around time with sample to reporting in 4 days, is very high-resolution and has the ability to detect all SV classes in an automated fashion.

## Data Availability

All data has been made available in the manuscript

## Acknowledgments

The authors thank Drs. Jim Broach, Viola Alesi, and Fariborz Rashid-Kolvear, for providing de-identified samples and Henry Sadowski for input to optimize the prenatal assay workflow. We thank Disorders of Sex Development Translational Research Network funded by NICHD (grant# R01HD093450) for providing the diagnostic results for 49,XXXXY case.

